# Age and Task-Dependent Modulations in EMG-EMG Coherence during Gait: A Scoping Review

**DOI:** 10.1101/2025.03.14.25323927

**Authors:** Silvère De Freitas, Fabien Dal Maso, Laetitia Fradet, Denis Arvisais, Yosra Cherni

## Abstract

Based on electromyography (EMG) recordings, EMG-EMG coherence method provides a practical approach to investigate neural mechanisms involved in locomotion. Although some studies indicated an influence of age and walking conditions on EMG-EMG coherence, no clear consensus emerged from the existing literature. The aim of this scoping review was to map the current literature on EMG-EMG coherence in healthy adults across ages and walking tasks. Six databases (CINAHL, Cochrane Central, CDSR, MEDLINE, Embase, and Web of Science) were searched, resulting in 31 studies included (575 healthy individuals). These studies analyzed EMG-EMG coherence of muscles involved during different locomotor tasks. The results revealed a consensus regarding the decrease in EMG-EMG coherence during walking with aging, particularly in the Beta and Gamma bands, which could be attributed to natural alterations in the corticospinal tract with age. Furthermore, Beta and Gamma EMG-EMG coherence showed an increased tendency during challenging proprioceptive and proactive locomotor tasks, which is interpreted as an enhancement of cortical involvement in gait control. This review also highlights the necessity for future research to examine EMG-EMG coherence in additional frequency bands, such as Alpha, utilizing standardized signal processing techniques and frequency classifications, and to investigate coherence in children across various locomotor tasks.

**NEW & NOTEWORTHY:** The EMG-EMG coherence method, based on electromyography recordings, is a practical tool to study neural mechanisms in locomotion. This scoping review explores the effect of age and walking conditions on EMG-EMG coherence during gait in healthy individuals. The results revealed a strong tendency of EMG-EMG coherence to decrease during aging and to increase during challenging proprioceptive and proactive locomotor tasks. This review also highlights gaps knowledge in children, and methodological concerns for coherence assessment.

## INTRODUCTION

Human locomotion is central to most daily living activities, that require navigating challenging environments such as for instance traversing uneven surfaces, overcoming obstacles to reach a destination. While often perceived as a simple and automatic motor task, walking is a complex task that results from dynamic interactions between cortical and spinal neural networks to coordinate body segment and maintain stability (1, 2). Neural activity in the spinal cord is thought to be driven by central pattern generators (3), but the contribution of sensory and supra-spinal control appears to be crucial for adapting the gait pattern to environmental constraints (4, 5). Thus, the neuromotor control of human locomotion remains a key area of investigation.

To explore the neural control of human locomotion, EMG-EMG (electromyography) coherence represents a promising approach. Coherence measures the coupling between oscillatory activity between two signals in the frequency domain (6–8) and can be assessed by measuring a pair of EMG signals non-invasively. Common oscillatory inputs are quantified between pairs of muscles (*i.e.,* inter-muscular coherence), within different parts of the same muscle (*i.e.,* intra-muscular coherence), for a given frequency band (9). EMG-EMG coherence observed frequency bands described thereafter is thought to originate from distinct brain regions and neural circuits (*i.e.,* cortical, subcortical, spinal) (10, 11). The Delta band (0-5 Hz) supports low-level motor control by synchronizing motor unit firing and can be attributed to subcortical activity (10, 12). The Alpha band (∼10 Hz) reflects subcortical and some cortical inputs, required to adjust postural control during locomotion (10, 13). The Beta band (∼20 Hz) is associated with corticospinal drive to maintain gait control (9, 14, 15). The gamma band (>30Hz) appears to be linked to corticospinal-originating signals (16) with an involvement in fine muscular coordination and adaptation to changing environments during human walking (17). Literature sometimes mentions a frequency range of 30-60 Hz, known as the Piper frequency band, which overlaps with the gamma frequency band (16). Although, the exact neurophysiological mechanisms underlying EMG-EMG coherence in those frequency bands vary across different sources in the literature, EMG–EMG coherence in the beta to low-gamma frequency bands emerged as a valuable marker of the contribution of the neuromotor networks involved during walking (15, 18, 19).

The current literature underscores significant variations in cortical dynamics across different temporal and spectral patterns, influenced by the diverse range of walking tasks in healthy individuals (*e.g.,* treadmill walking at varying speeds, with stability challenges, or using robotic assistance) (20–22). According to these findings, EMG-EMG coherence in the Beta and Gamma bands increases for lower limb muscles during split-belt treadmill walking tasks (23), but not for the Gamma band during a fast walking task (16), highlighting the particular role of the corticospinal tract in proprioceptive-driven locomotor adaptation and the specificity of EMG-EMG coherence response to task demands. In addition to walking tasks, muscle coherence seems to be modulated by age during walking (24, 25). In childhood development (4–15 years), EMG-EMG coherence in the Piper frequency band within the tibialis anterior (TA) increases and is associated with improved ankle control, as shown by reduced toe position variability during swing phase (16). Then, aging may induce functional changes in the corticospinal tract to the ankle muscles, as shown by a distinct response in older adults, with reduced Gamma band inter-muscular coherence compared to younger adults who show no changes during dual-task (cognitive-motor) (16, 26). However, target walking tasks show similar responses, such as an increase in intra-muscular coherence, in both young and elderly adults (27, 28). Therefore, the impact of maturation and aging on EMG-EMG coherence during walking, as well as the influence of challenging task conditions, remains unclear, with no consensus in the current literature.

This scoping review aims to (i) summarize research on the effect of age and locomotor tasks on EMG-EMG coherence in healthy individuals; (ii) report the different methodological approaches for EMG-EMG coherence analysis.

## MATERIALS AND METHODS

### Protocol and search strategies

This scoping review was conducted according to the Preferred Reporting Items for Systematic reviews and Meta-Analyses (PRISMA) adapted for Scoping Reviews checklist (29). We searched in Ovid MEDLINE, Ovid EMBASE, Ovid Cochrane Central Register of Controlled Trials, Ovid Cochrane Database of Systematic Reviews, EBSCO Cinahl Complete and Clarivate Web of Science Core Collection. A comprehensive literature search was conducted by an experienced librarian (DA) first in the MEDLINE database and then translated to the other databases. The search strategy used a combination of descriptors and keywords. Search terms were related to muscle coherences, lower limbs, and gait. There were no language restrictions. The search was last conducted on March 29, 2024. The search strategy for Medline can be found in the Appendix 1.

### Study selection

The inclusion criteria were: (a) peer-reviewed experimental studies, observational studies, and clinical trials, excluding reviews, editorials, and case reports; (b) studies including healthy human participants of any age group, for studies focusing on gait-related pathologies, only the control group was considered, if applicable; (c) studies where the primary outcome was EMG-EMG coherence during gait.

### Study screening

First, all identified references were compiled and uploaded to the Covidence software (Melbourne, Victoria, Australia) to remove duplicates and initiate the source selection process. Second, titles and abstracts were screened independently by two evaluators (S.T. and S.E.), based on inclusion criteria. Third, the selected articles were full-text screened by two authors (S.T. and Y.C.). In cases of disagreement between the two authors, the opinion of a third evaluator (F.D.M.) was considered to determine final inclusion.

### Data Charting Process and Analysis

Results were organized and compiled into tables and charts. The general information were presented based on: (1) bibliographic characteristics (authors and year); (2) sample characteristics (number, age and sex); (3) task categories (classification of tasks based on their functional demands); (4) locomotor task (walking task performed under specific conditions, *e.g.,* surface, walking speed, modality); (5) methodology of coherence analysis (number of gait studied, muscles recorded, EMG pre-processing method, power spectrum computation, and frequency band studied); (6) main findings regarding the effects of age and locomotor tasks EMG-EMG coherence. Effect sizes (ES) of each statistically significant main effect (p<0.05) of age, locomotor task (*vs*. regular walk), and condition on intra- and inter-muscular coherence were reported in four frequency bands (Delta, Alpha, Beta, Gamma), where applicable. For studies that did not report the ES, the latter was calculated from the mean and standard deviation provided in the articles, if applicable. The authors were contacted if none of that information was available. For the studies that used parametric tests, a normal distribution of the data was assumed, and a Cohen’s d ES (*d*) was calculated (30). For non-parametric tests, a Glass’s delta ES (△) was calculated (31). Effect size below 0.2 were considered as very small, 0.2–0.5 as small, 0.5–0.8 as medium, 0.8–1.0 as large, and those above 1.0 as very large (32). Data extraction was conducted by the first author (S.D.) and validated by two other authors (Y.C. & F.D.M.).

## RESULTS

### Search Results

The initial databases search yielded a total of 6,399 articles. After removing duplicate articles (n = 2,819), 3,580 articles were screened for title and abstract. Then, 249 articles were assessed for eligibility by full-text screening, leading to the inclusion of 31 studies. The flowchart of the selection process is shown in Figure 1.

**Figure 1:**
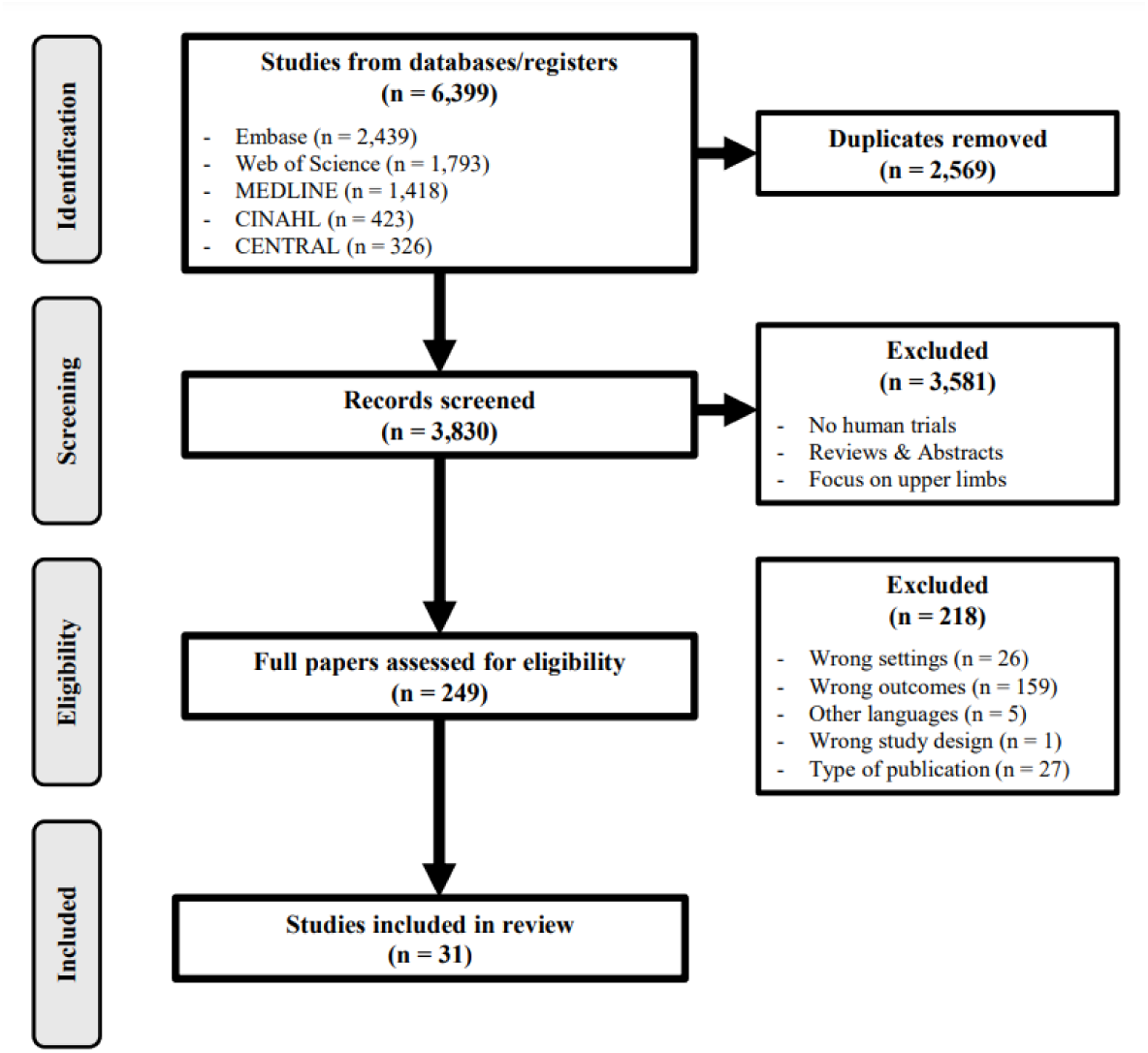
PRISMA flowchart of the selection process

### Studies characteristics

Included studies were published between 2003 and 2024. A total of 575 healthy participants (age ranging from 10 months to 75 years old) were included. An overall view of age groups, muscles and frequency bands studied, locomotor tasks, and task categories of the included studies is illustrated in Figure 2.

**Figure 2:**
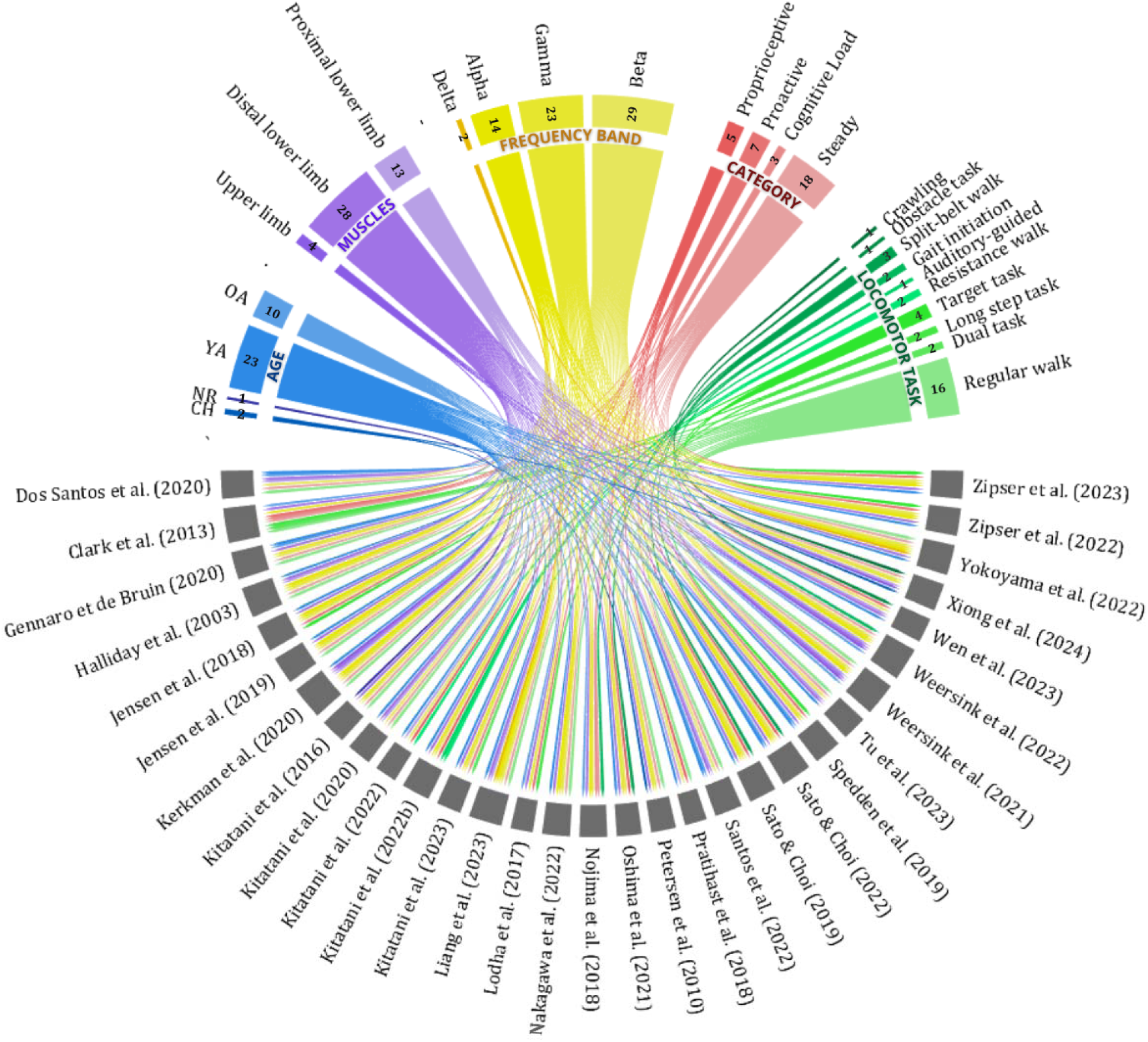
Distribution of Age, group of Muscles, Frequency band, Task and Category of task studied among the included studies. Abbreviations: children (CH); old adult (OA); young adult (YA); not reported (NR).

### Walking tasks

All tasks were classified into five main categories of walking tasks performed by the participants: “Steady state” (no external perturbations), “Proactive” (anticipatory actions), “Proprioceptive” (actions based on internal sensory feedback, *i.e.* afferent signal from proprioceptive sensor responding to variations in position, movement and balance), and “Cognitive Load” (additional cognitive demands) (Figure 2). Eighteen studies (58%) analyzed steady state walking tasks, including regular walking at various speeds, physiological states, and different locomotion patterns (*e.g.,* crawling and gait initiation). Seven studies (23%) investigated proactive tasks, such as obstacles, targets, long step, and auditory guided walking task. Five studies (16%) examined proprioceptive tasks, involving split belt, unilateral resistance, and high heeled shoes walking tasks. And three studies analyzed additional cognitive load (10%), such as auditory 2-back task, watching figures, digit 2-back task during a walking task and choice reaction task during gait initiation. Among these tasks, participants walked on a treadmill in 23 studies (74%) and on the ground in 8 studies (26%).

### Methodology

Table 1 presents the methodological characteristics of the 31 included studies. (1) bibliographic characteristics (authors and year); (2) sample characteristics (number, age and sex); (3) locomotor task (walking task performed under specific conditions); (4) methodology of coherence analysis (number of gait studied, muscles pairs recorded, gait phase of interest, EMG pre-processing method, power spectrum computation, and frequency band studied).

**Table 1:**
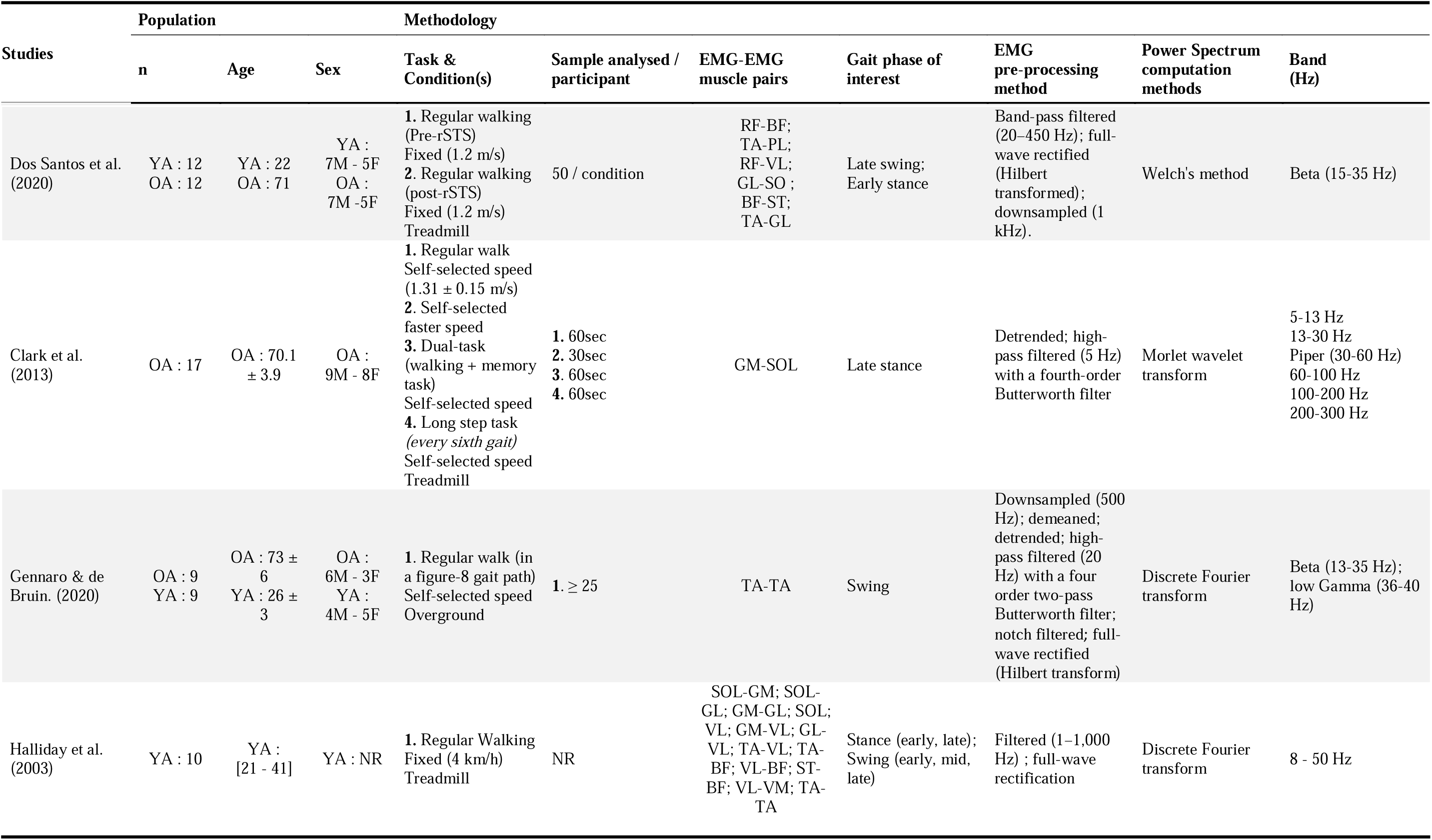
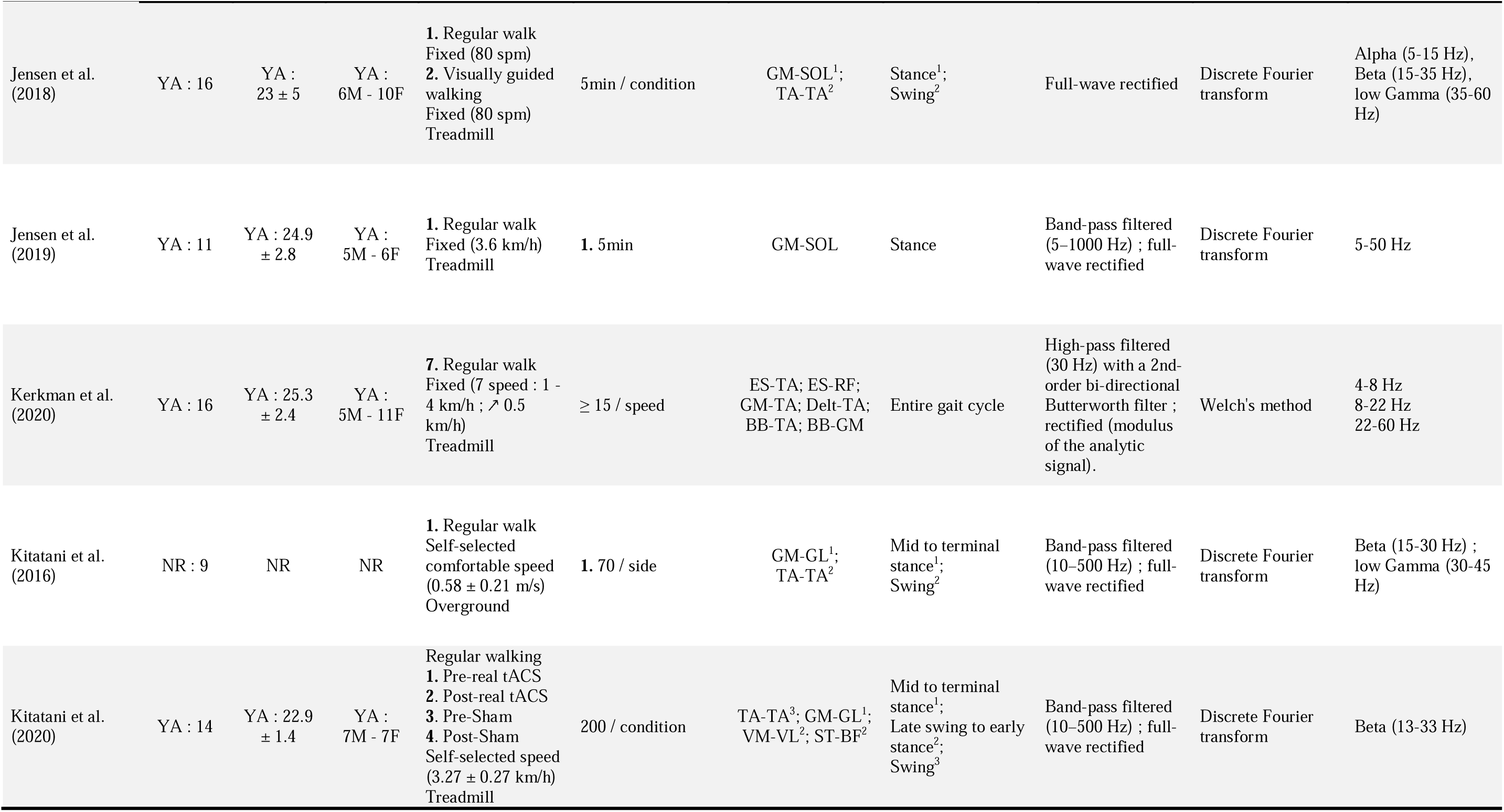
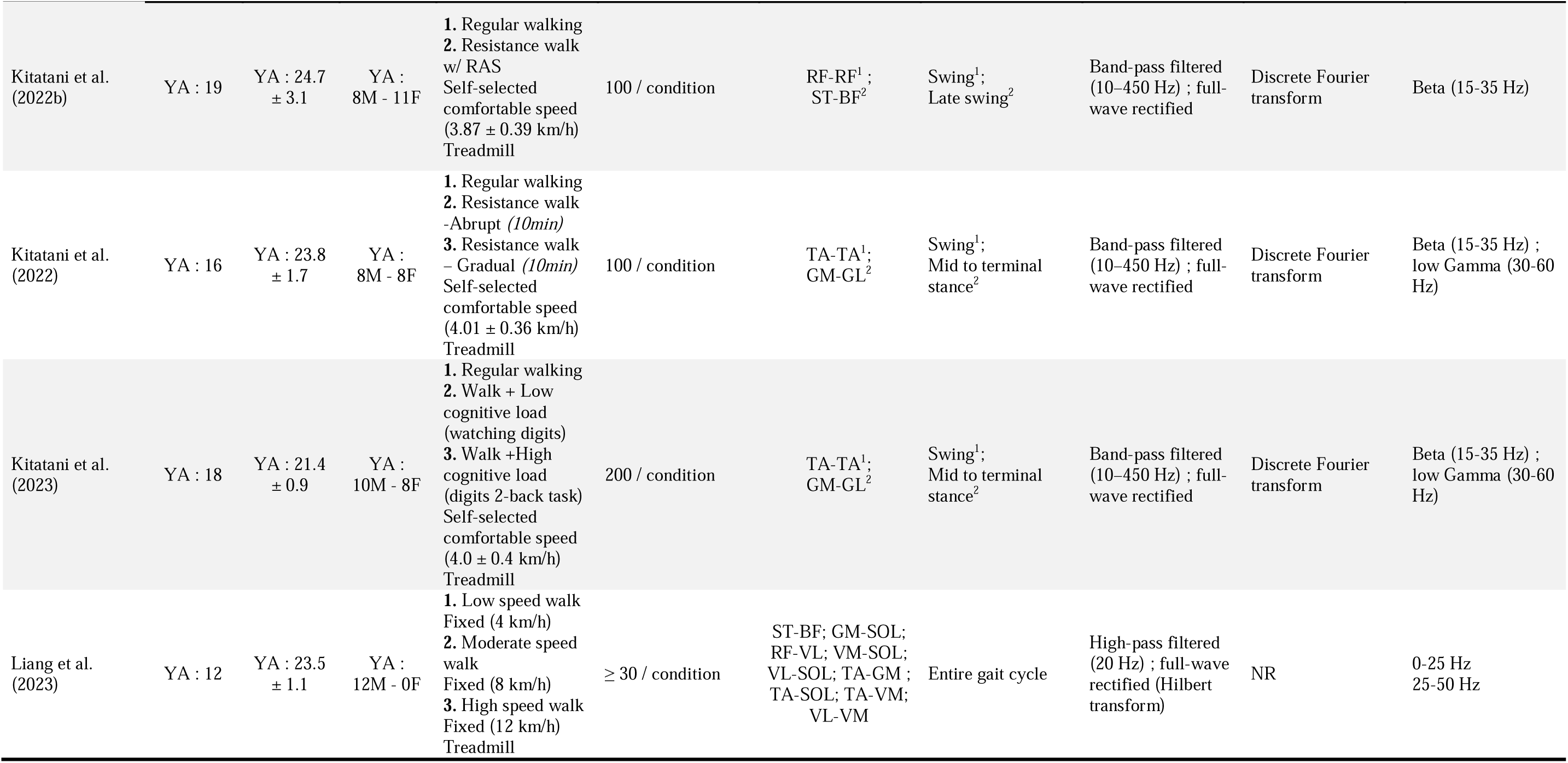
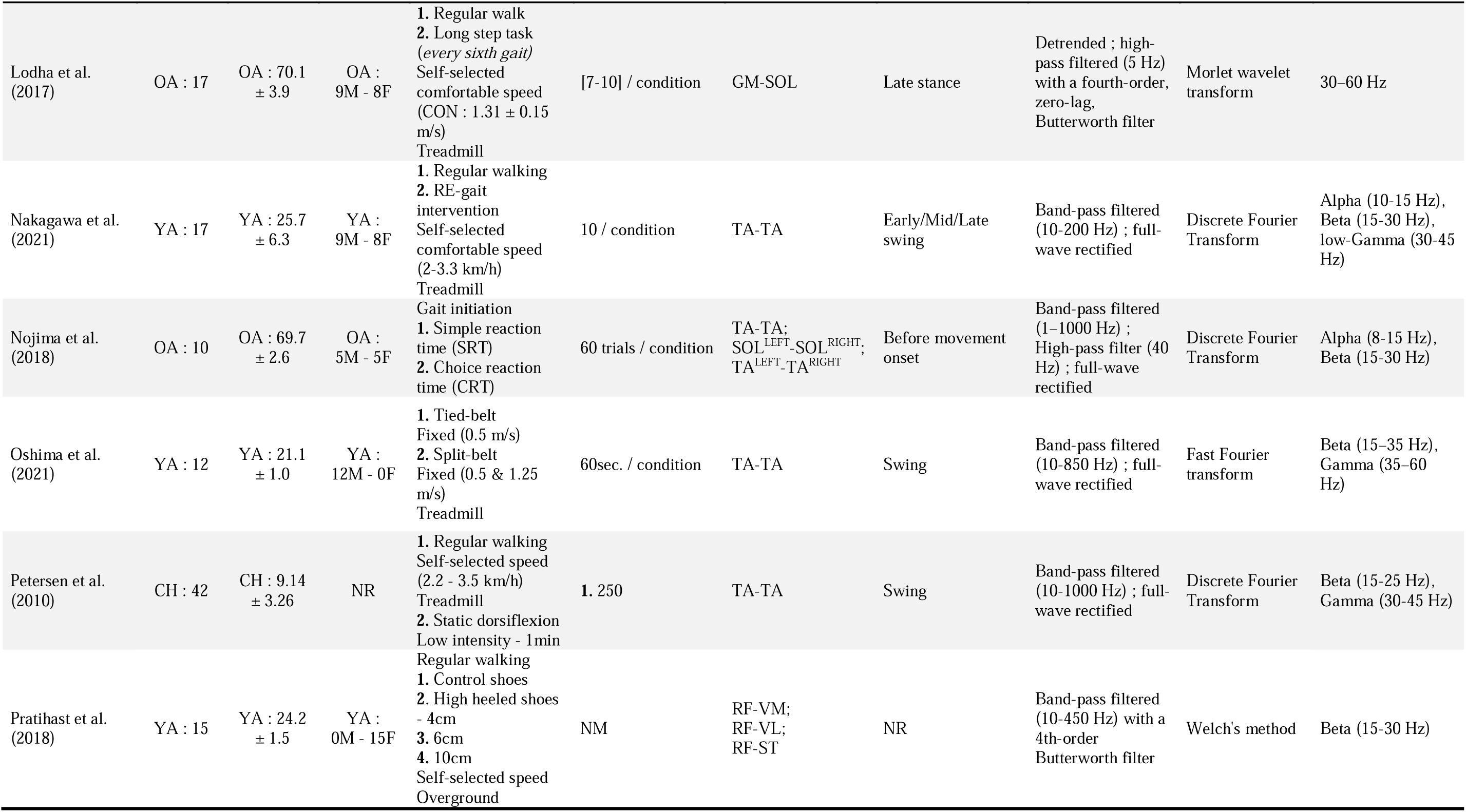
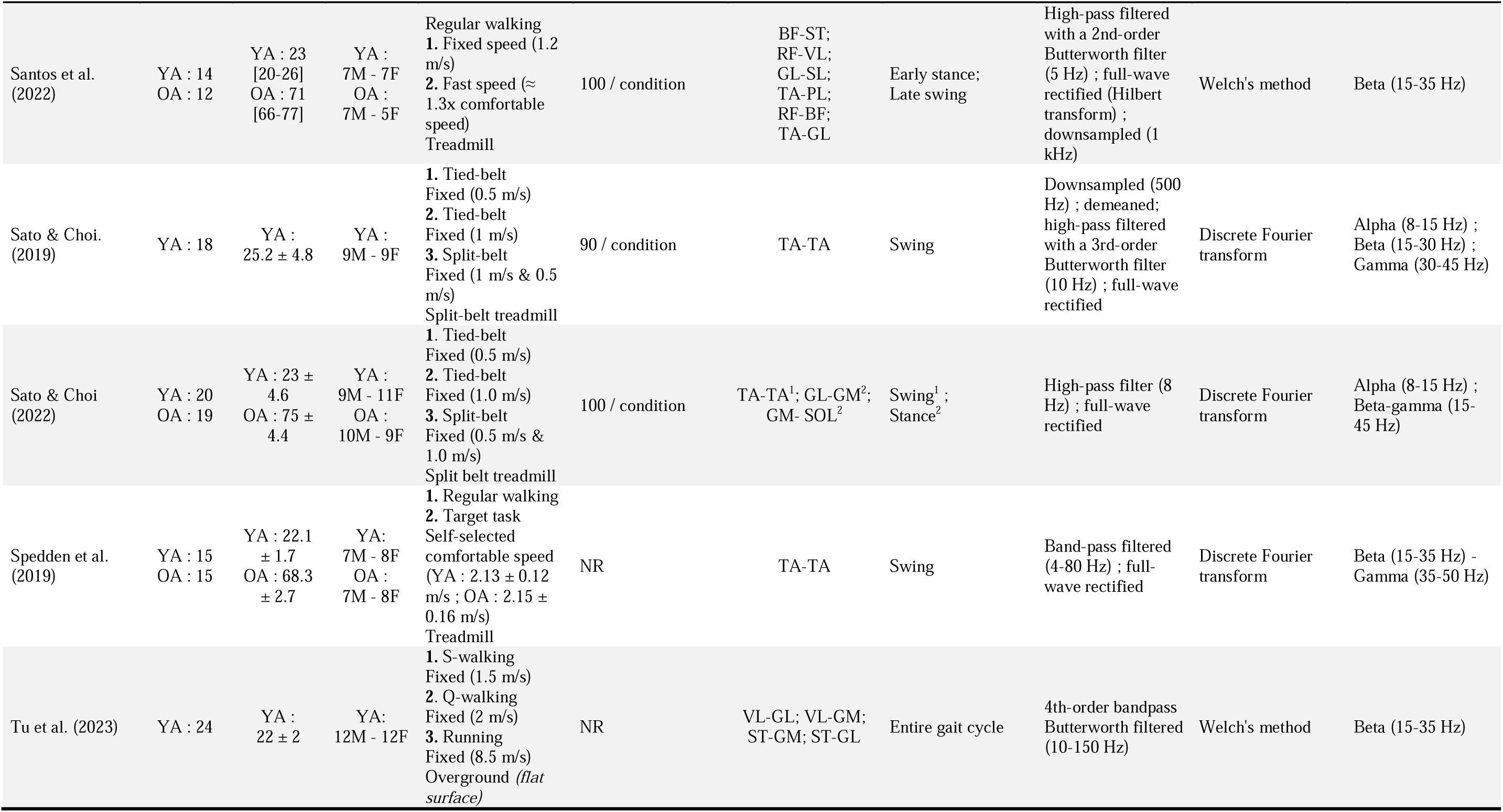
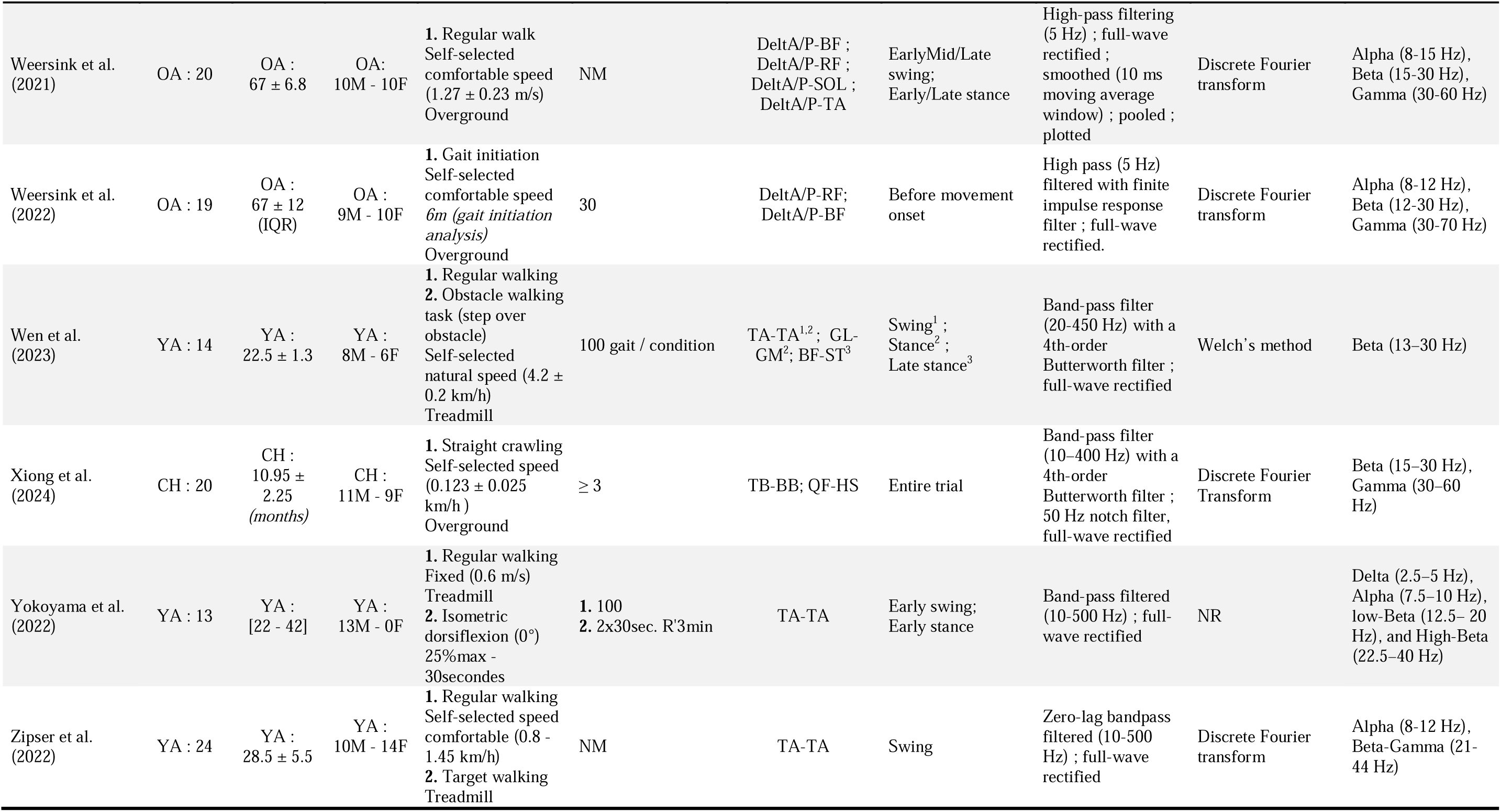
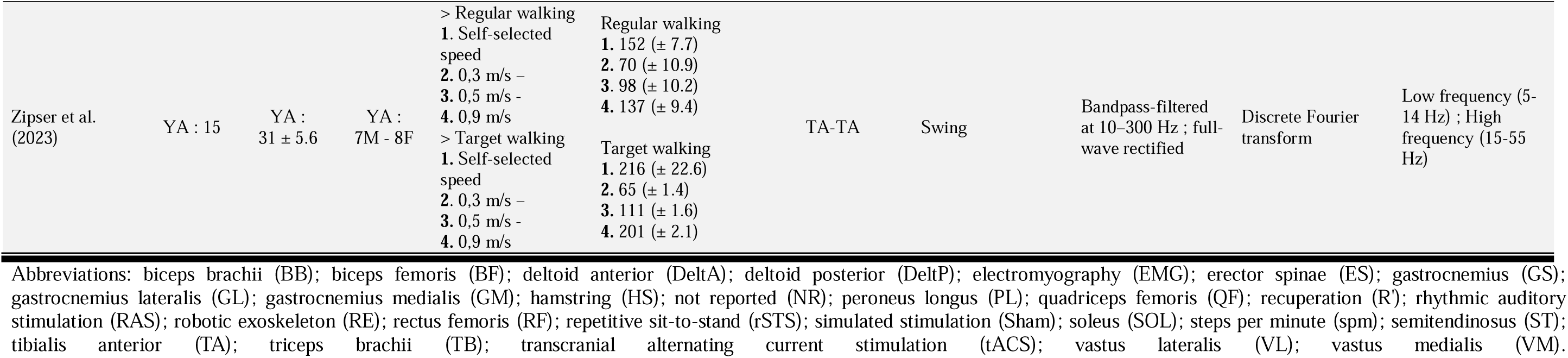
Population characteristics and methodological information of the included studies.

#### Population

The studies’ sample size ranged from 9 to 42 participants (mean ± SD of 19 ± 8). Children were 6.53 ± 4.70 years old, young adults were 24.68 ± 3.01 years old, and old adults 70.22 ± 2.38 years old (Table 1). Young adults were the most studied age group in the included studies (23 studies, total of 354 participants), followed by old adults (10 studies, 150 participants), and finally children (two studies, 62 participants) (see Figure 2). One study did not report the exact age range of participants (9 old adults) (33).

#### Muscles and muscle pairs

Most studies (28 studies - 90%) assessed EMG-EMG coherence using distal lower limb muscles, 13 studies (42%) included proximal lower limb muscles, and 4 studies (13%) upper limbs and proximal/distal lower limbs (Figure 2, Table 1). Nineteen studies (61%) investigated intra-muscular coherence of which 18 studies measured intra-muscular coherence within the TA and one between proximal and distal portions of the rectus femoris (34), 22 studies (71%) investigated inter-muscular coherence, and 10 studies (32%) investigated both intra- and inter-muscular coherence. Inter-muscular coherence was measured over a large panel of muscles, distributed between lower and upper limbs (Table 1). Distal lower limb muscles were the most studied during locomotor tasks (Figure 3). TA was the most studied muscle (23 studies – 74% of total studies) followed gastrocnemius medialis (GM), gastrocnemius lateralis (GL), soleus (SOL) (10-14 studies). Peroneus longus (PL) was studied only twice (35, 36). Proximal lower limb muscles (rectus femoris (RF), vastus medialis (VM), vastus lateralis (VL), biceps femoris (BF), semitendinosus (ST)) were studied in between 4 to 9 studies, with the highest representation from the knee flexors (BF, ST) and RF (9 and 8 studies, respectively), and the lowest from the VM (4 studies) (Figure 3, Table 1). The upper limb muscles were minimally (deltoid (Delt), biceps brachii (BB), triceps brachii (TB), erector spinae (ES)). The deltoid (including anterior and posterior heads) showed the highest representation, being included in 3 studies, followed by the BB in 2 studies, and the ES and TB in 1 study each (Figure 3, Table 1).

**Figure 3:**
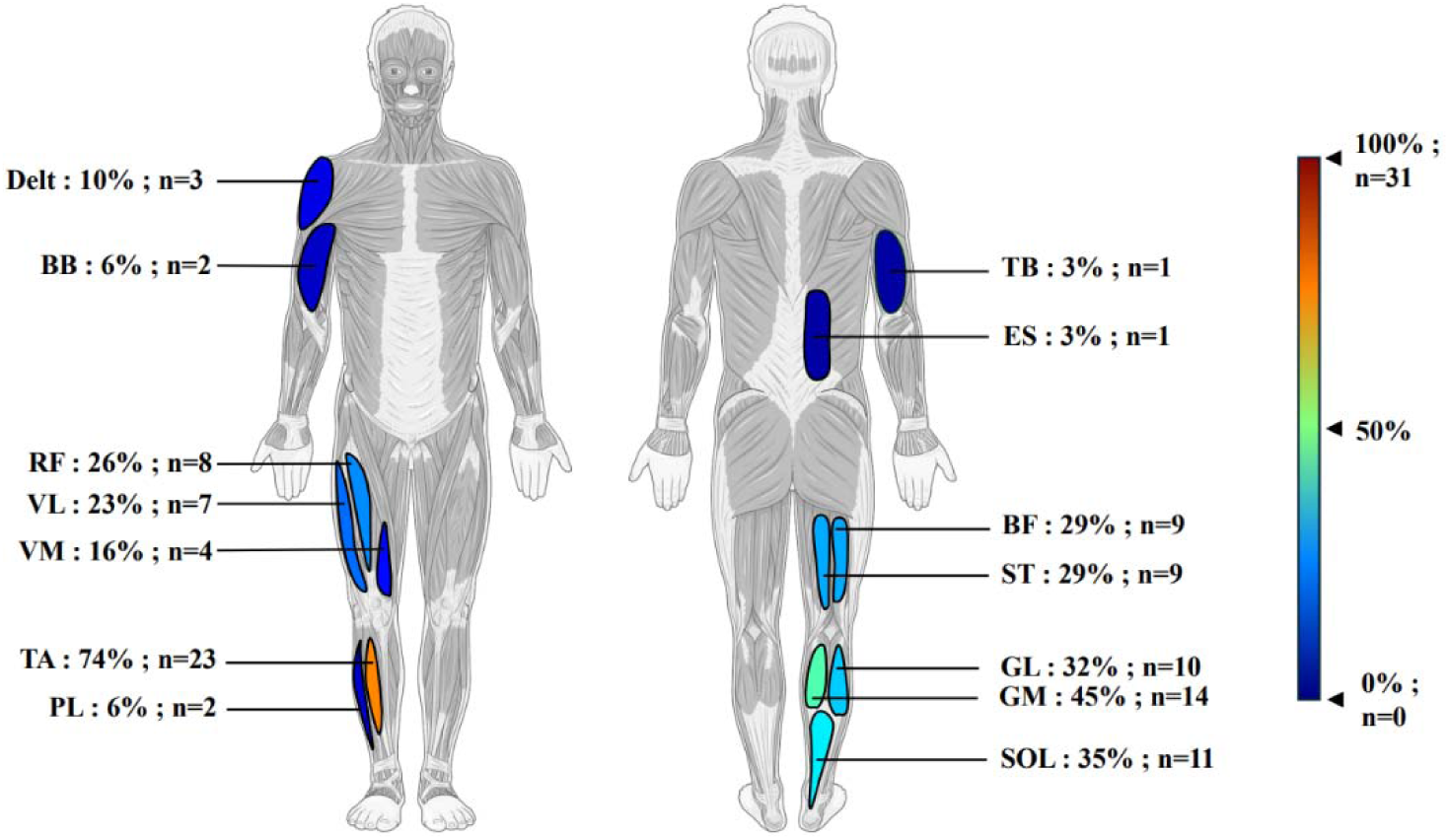
Distribution of muscles studied for EMG-EMG coherence during gait. Biceps brachii (BB); biceps femoris (BF); deltoid (Delt); erector spinae (ES); gastrocnemius lateralis (GL); gastrocnemius medialis (GM); number of studies (n); peroneus longus (PL); rectus femoris (RF); soleus (SOL); semitendinosus (ST); tibialis anterior (TA); triceps brachii (TB); vastus lateralis (VL); vastus medialis (VM). *This figure were drawn using images from Servier Medical Art. Servier Medical Art by Servier is licensed under a Creative Commons Attribution 4.0 Unported License (CC BY) (*https://smart.servier.com/citation-sharing/*) (*https://creativecommons.org/licenses/by/4.0/).

#### EMG pre-processing

Twenty studies (65%) band-pass filtered EMG signals with a low cut-off frequency ranging from 4 Hz to 20 Hz and a high cut-off frequency ranging from 80 Hz to 1,000 Hz (Table 1). Ten studies (32%) high-pass filtered EMG signals with a cut-off frequency ranging from 5 Hz to 40 Hz. One study did not report a filtering EMG signals (27). Twenty seven studies (87%) full-wave rectified EMG signals before coherence computation, and four utilized the raw signal (16, 37–39).

#### Power Spectrum computation

Twenty-seven studies (87%) used Fourier-based methods (discrete Fourier transform, fast Fourier transform, Welch’s method) to compute power spectrum density (Table 1). Two studies used the Morlet wavelet transform (16, 38), and two did not specify the power spectrum computation method (40, 41).

#### Gait phase of interest

Most studies analyzed EMG-EMG coherence during specific gait phases and sub-phases (Table 1). Intra-muscular coherence (TA-TA and RF-RF) was assessed during the swing phase in all studies (19), with eight (42%) further dividing it into early, mid, and late swing. Only one study also examined TA-TA coherence during the stance phase (41). Inter-muscular coherence was analyzed during both swing and stance phases, with sub-phase decomposition in 13 studies (59%). Four studies considered the entire gait cycle (39, 40, 42, 43), two studies focused on gait initiation tasks (21, 44), and one study did not specify (37) (Table 1).

#### Frequency band

Studies measured EMG-EMG coherence in the Delta, Alpha, Beta, and Gamma bands during walking. However, the frequency range for each frequency band varied between selected studies (Figure 4). Delta band was investigated only once and identifying by a 2.5-5 Hz range (41). Nine studies out of 14 (64%) selected 8–12 Hz for the coherence analysis of the Alpha band (Figure 3B), 21 studies out of 29 selected 15–30 Hz for the Beta band (72%) (Figure 3C), and nine to 15 studies out of 23 selected 30-60 Hz for the Gamma band (39 to 65%) (Figure 3D). An overlap is observed between the Alpha and Beta bands in the range of 12 to 15 Hz, as well as between the Beta and Gamma bands in the range of 30 to 40 Hz (Figure 3A). Two studies combined the Beta and Gamma frequency bands (45, 46). For EMG-EMG coherence analysis during walking, the Beta band was the most analyzed frequency band (29/31 – 94% studies) (Figure 2), two studies analyzed only Gamma frequency band (16, 38). Five studies have also investigated larger frequency band without classifying them in the frequency bands previously cited (*e.g.,* Low/High frequency band) (14, 40, 42, 47, 48).

**Figure 4:**
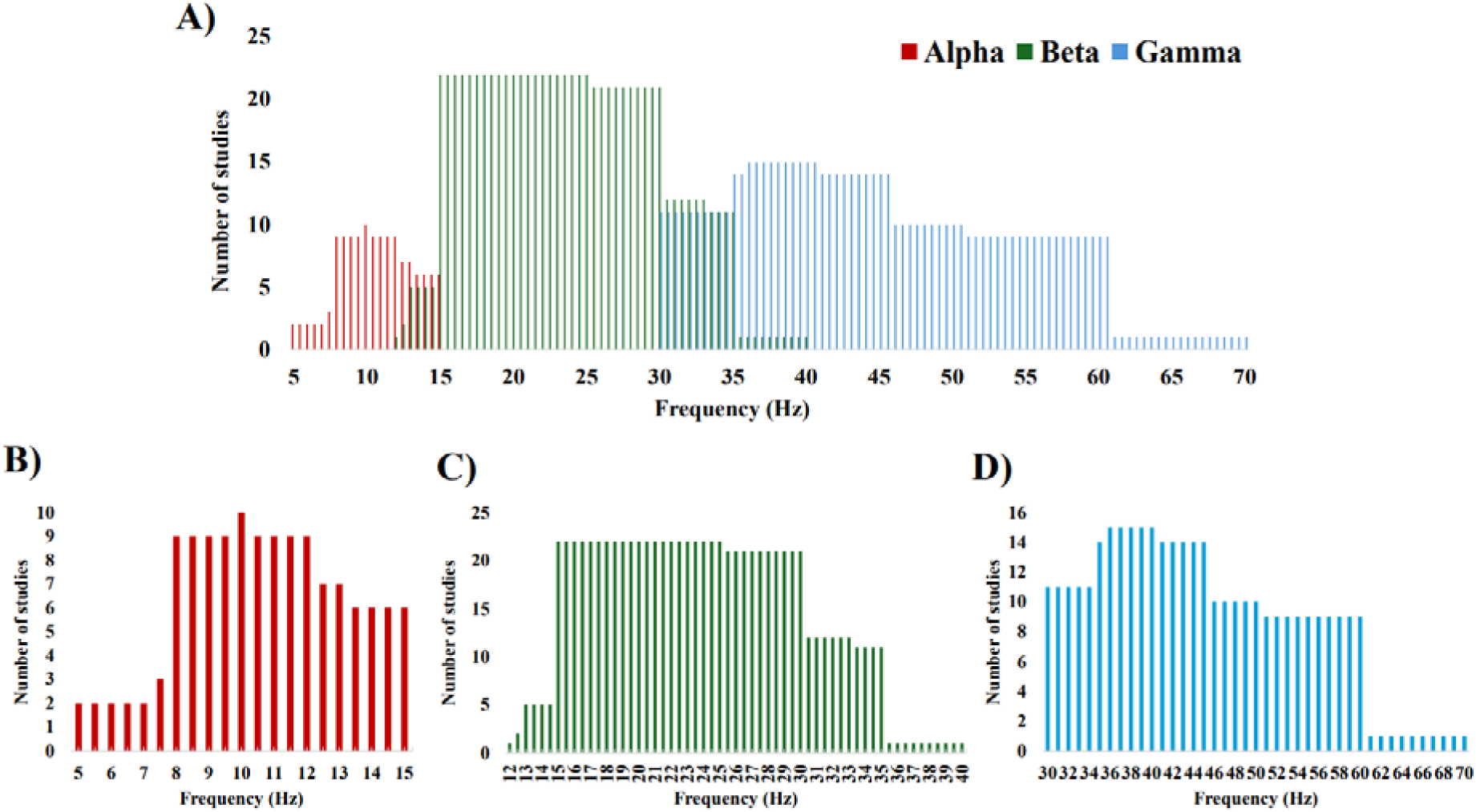
Number of studies that included each frequency in the Alpha, Beta, and Gamma bands. **A)** Alpha, Beta and Gamma band; **B)** Alpha band; **C)** Beta band; **D)** Gamma band.

### Effect of age on EMG-EMG coherence

#### Children

One study was conducted to quantify the difference between different age groups of children in TA-TA coherence during walking (24). EMG-EMG coherence in the Gamma band significantly increases with maturation while walking (24). Children aged 7-9, 10-12 and 13-15 years showed a higher coherence in the Gamma band compared to younger 4-6 years children during a regular walking task executed in a steady state manner (Table 3). A moderate significant positive correlation was shown between age and peak Gamma band coherence during walking (r = 0.54) (24). Gamma coherence was observed between TB and BB in crawling infants with typical development, no comparison with other age groups was made (43).

#### Old adults vs young adults

All studies (5 studies) that investigated the effect of age between old adults and young adults on EMG-EMG coherence consistently showed lower coherence in older adults than their younger counterparts (Table 2). This was evidenced in the Beta and Gamma bands TA-TA coherence during regular walking (ES = 1.55, large effect) (49), as well as inter-muscular coherence in synergistic lower limb muscles (TA-PL, RF-VL, GL-SOL) for the Beta band (ES = 1.21–1.41, large effect) (35, 36) (Table 2). In fast walking, older adults shown lower TA-PL and GL-SOL coherence than younger adults in Beta band (36) (Table 2). Under muscle fatigue (*e.g*., repetitive sit-to-stand), older adults showed lower RF-VL (ES = 0.86, large effect) and TA-PL coherences (ES = 0.76, medium effect) than younger adults in the Beta band (35). In target walking tasks, age effects were larger in late swing for Beta (ES = 0.47, η²p) and Gamma (ES = 0.38, η²p) bands compared to early swing (Beta: ES = 0.16, η²p; Gamma: ES = 0.19, η²p) (28). In split-belt walking tasks (asymmetrical walking speeds) (45), older adults had lower TA-TA coherence in Alpha (ES = 0.51–0.53, medium effect) and Beta-Gamma bands (ES = 0.70– 0.81, medium/large effect). Inter-muscular coherence in distal synergistic muscles (GL-GM, GM-SOL) was also lower in older adults in Alpha (ES = 0.41–0.53) and Beta-Gamma (ES = 0.61– 0.71) bands (Table 2). No study has compared the Delta band between these two age groups during walking. One study reported no significant age-related difference in antagonist RF-BF muscles pair during the swing phase in regular walking (35) (Table 2), and different fatigue responses between age groups: GL-SOL coherence decreased in young adults (ES = 0.43, small effect) but didn’t significantly increase in older adults, with no significant group difference during walking (35) (Table 3).

**Table 2:**
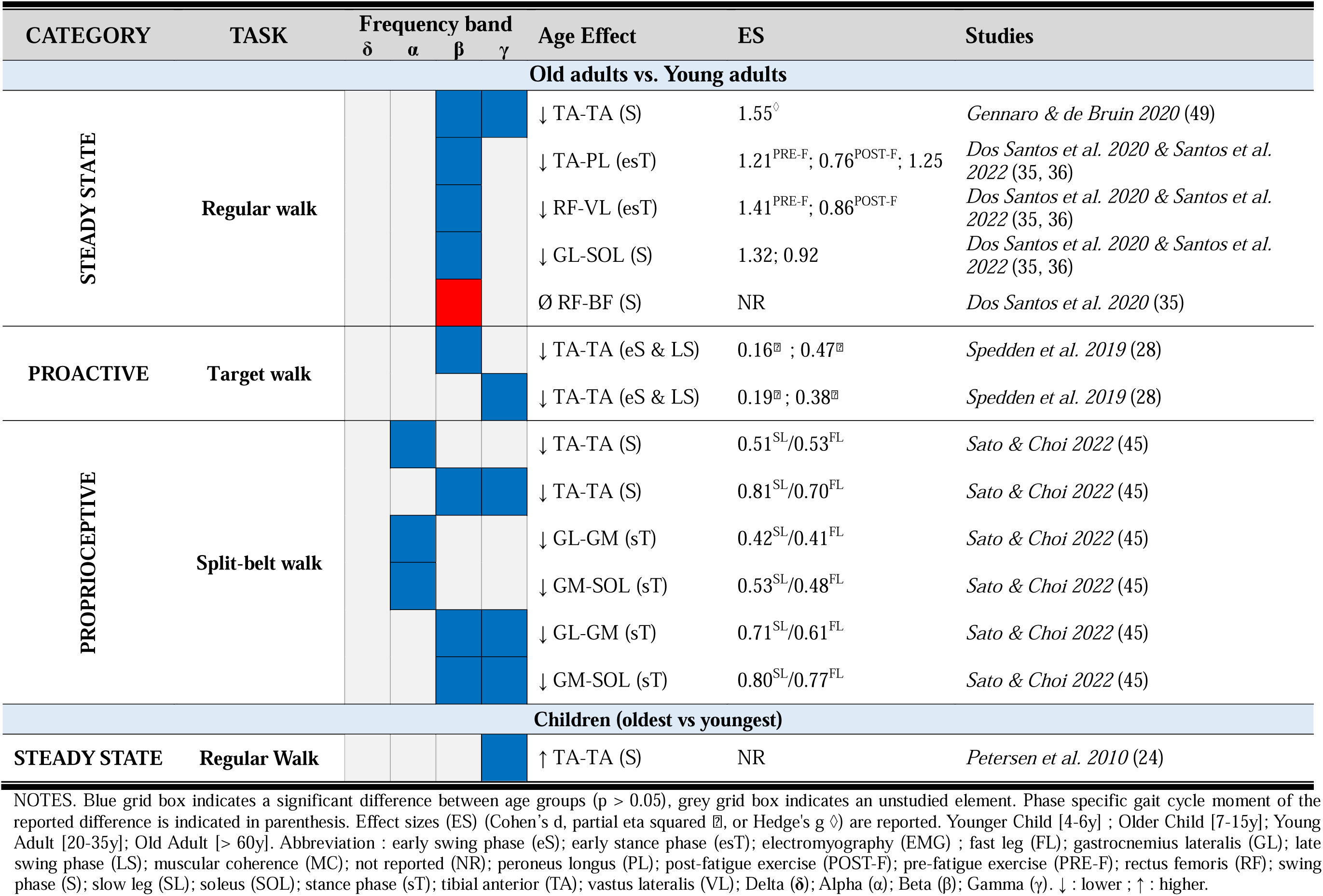
Effect of age on EMG-EMG coherence across different walking tasks and conditions for Delta, Alpha, Beta, and Gamma band.

| CATEGORY | TASK | Frequency band |  |  |  | Age Effect | ES | Studies |
| --- | --- | --- | --- | --- | --- | --- | --- | --- |
| | | $\delta$ | $\alpha$ | $\beta$ | $\gamma$ | | | |
| Old adults vs. Young adults |  |  |  |  |  |  |  |  |
| STEADY STATE | Regular walk |  |  |  |  | ↓ TA-TA (S) | 1.55<sup>◇</sup> | Gennaro & de Bruin 2020 (49) |
|  |  |  |  |  |  | ↓ TA-PL (esT) | 1.21<sup>PRE-F</sup>; 0.76<sup>POST-F</sup>; 1.25 | Dos Santos et al. 2020 & Santos et al. 2022 (35, 36) |
|  |  |  |  |  |  | ↓ RF-VL (esT) | 1.41<sup>PRE-F</sup>; 0.86<sup>POST-F</sup> | Dos Santos et al. 2020 & Santos et al. 2022 (35, 36) |
|  |  |  |  |  |  | ↓ GL-SOL (S) | 1.32; 0.92 | Dos Santos et al. 2020 & Santos et al. 2022 (35, 36) |
|  |  |  |  |  |  | Ø RF-BF (S) | NR | Dos Santos et al. 2020 (35) |
| PROACTIVE | Target walk |  |  |  |  | ↓ TA-TA (eS & LS) | 0.16<sup>▣</sup> ; 0.47<sup>▣</sup> | Spedden et al. 2019 (28) |
|  |  |  |  |  |  | ↓ TA-TA (eS & LS) | 0.19<sup>▣</sup> ; 0.38<sup>▣</sup> | Spedden et al. 2019 (28) |
| PROPRIOCEPTIVE | Split-belt walk |  |  |  |  | ↓ TA-TA (S) | 0.51<sup>SL</sup>/0.53<sup>FL</sup> | Sato & Choi 2022 (45) |
|  |  |  |  |  |  | ↓ TA-TA (S) | 0.81<sup>SL</sup>/0.70<sup>FL</sup> | Sato & Choi 2022 (45) |
|  |  |  |  |  |  | ↓ GL-GM (sT) | 0.42<sup>SL</sup>/0.41<sup>FL</sup> | Sato & Choi 2022 (45) |
|  |  |  |  |  |  | ↓ GM-SOL (sT) | 0.53<sup>SL</sup>/0.48<sup>FL</sup> | Sato & Choi 2022 (45) |
|  |  |  |  |  |  | ↓ GL-GM (sT) | 0.71<sup>SL</sup>/0.61<sup>FL</sup> | Sato & Choi 2022 (45) |
|  |  |  |  |  |  | ↓ GM-SOL (sT) | 0.80<sup>SL</sup>/0.77<sup>FL</sup> | Sato & Choi 2022 (45) |
| Children (oldest vs youngest) |  |  |  |  |  |  |  |  |
| STEADY STATE | Regular Walk |  |  |  |  | ↑ TA-TA (S) | NR | Petersen et al. 2010 (24) |

NOTES. Blue grid box indicates a significant difference between age groups ( $p > 0.05$ ), grey grid box indicates an unstudied element. Phase specific gait cycle moment of the reported difference is indicated in parenthesis. Effect sizes (ES) (Cohen's d, partial eta squared □ , or Hedge's g $\diamond$ ) are reported. Younger Child [4-6y] ; Older Child [7-15y]; Young Adult [20-35y]; Old Adult [ $> 60y$ ]. Abbreviation : early swing phase (eS); early stance phase (esT); electromyography (EMG) ; fast leg (FL); gastrocnemius lateralis (GL); late
swing phase (LS); muscular coherence (MC); not reported (NR); peroneus longus (PL); post-fatigue exercise (POST-F); pre-fatigue exercise (PRE-F); rectus femoris (RF); swing phase (S); slow leg (SL); soleus (SOL); stance phase (sT); tibial anterior (TA); vastus lateralis (VL); Delta ( $\delta$ ); Alpha ( $\alpha$ ); Beta ( $\beta$ ); Gamma ( $\gamma$ ). ↓ : lower ; ↑ : higher.

### Effect of locomotor tasks on EMG-EMG coherence

#### Steady state

During steady walking at a comfortable speed, EMG-EMG coherence varied across gait phases and conditions. For young adults, intra-muscular coherence (TA-TA) increased during early stance (Delta, Beta) (41), and early/late swing (Alpha, Beta) (14) but decreased in mid-swing (14). No significant difference was observed between early stance and early swing in the Alpha band (41). Inter-muscular coherence increased in plantar flexors (GL-SOL, GM-SOL) across Alpha, Beta, and Gamma bands (14, 27) and in GM-GL during stance (14, 33). Low TA-GL coherence (>10 Hz, 0.015–0.065) was observed in early stance (33). Upper-limb muscles (anterior (DeltA)/posterior (DeltP) deltoids) showed increased coherence with proximal (BF, RF) and distal (SOL, TA) lower-limb muscles (50) (Table 3). During fast walking, TA-TA coherence increased in the Alpha and Beta band during swing (48) but, overall, showed no significant changes in Gamma (Table 3). Walking speed had a large effect on Beta coherence (ES = 0.26, η²p), with a 69% stronger TA-GL coherence during early stance (ES = 0.59, medium effect) for both age groups (36). Inter-muscular coherence between upper and lower limbs increased across all frequency bands according to one study (Delta to Gamma) (40). Fatigue from repetitive sit-to-stand tasks reduced proximal inter-muscular coherence (RF-BF; ES = 0.35, small effect) and increased distal coherence (TA-PL; ES = 0.21, small effect) in Beta band during swing and stance, respectively (35) (Table 3). Interventions like transcranial alternating current stimulation (51) and robotic ankle-assisted gait (20) increased swing-phase TA-TA coherence in Beta band, with a large effect of robotic exoskeleton (ES = 0.18, η²p), but had no effect in Alpha and Gamma bands (Table 3). During gait initiation, coherence increased between upper and lower limb muscles (DeltA/P-BF, DeltA/P-RF) across Alpha, Beta, and Gamma bands (44), as well as between distal lower limb muscles (TA-TA, TAprox_left_-TAprox_right_, SOL_left_-SOL_right_) in Alpha and Beta bands (21) (Table 3).

**Table 3:**
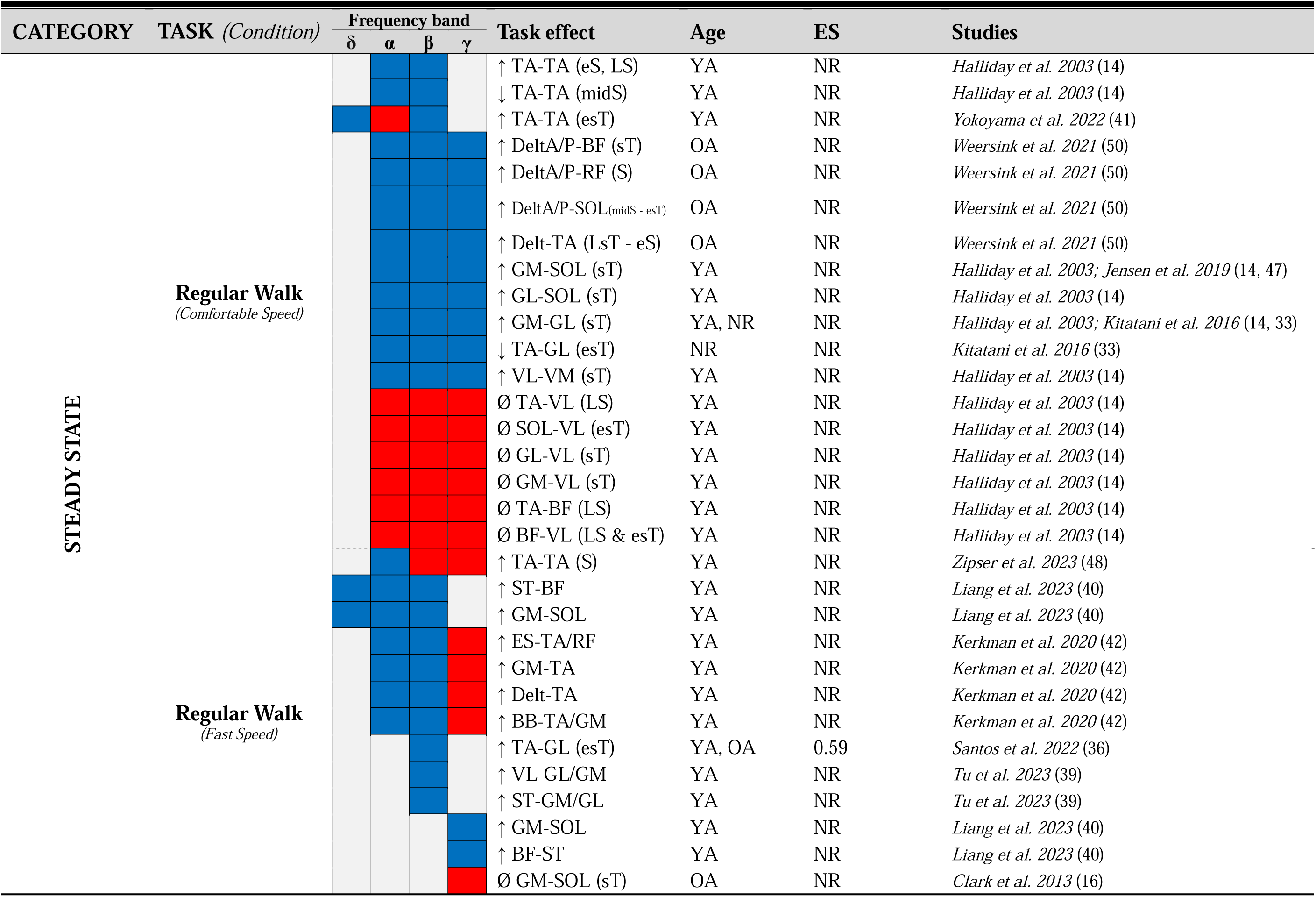
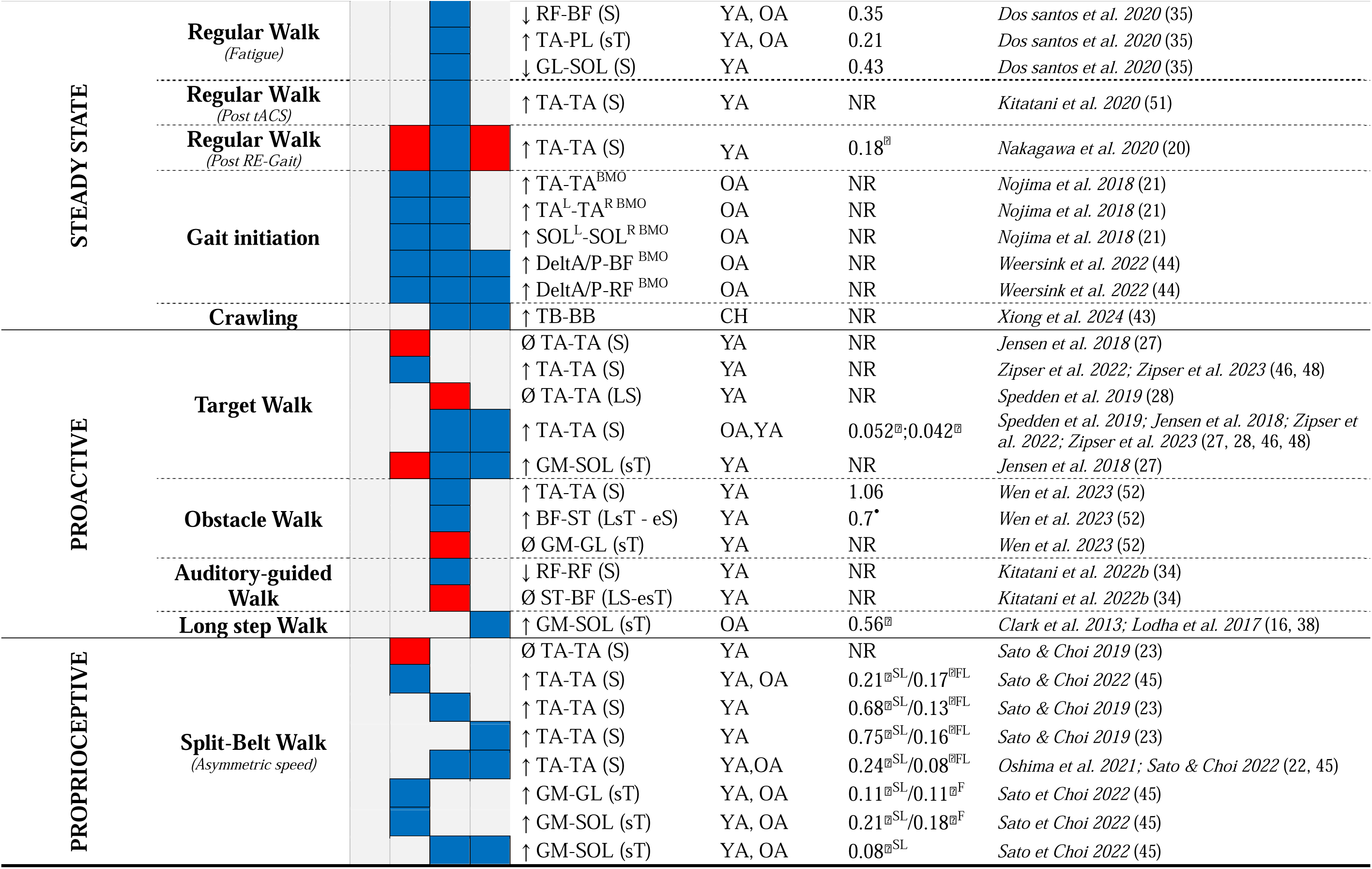
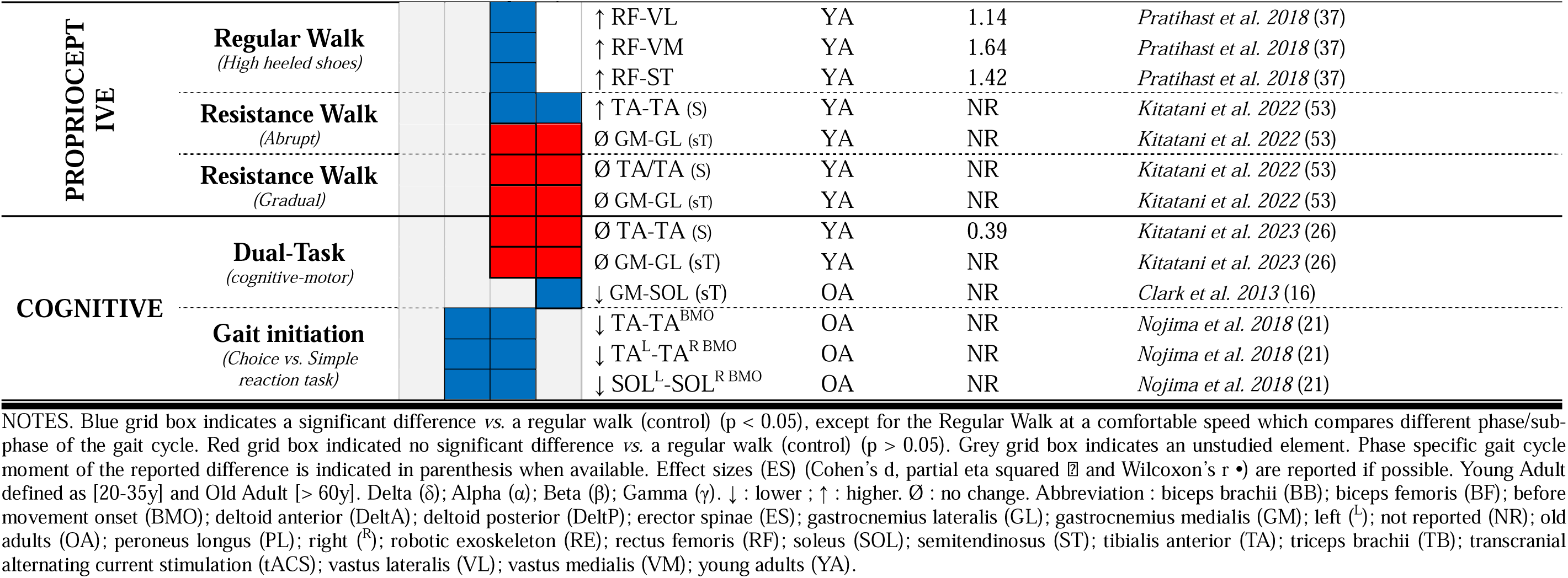
Effect of different locomotor tasks on EMG-EMG coherence across different frequency bands and age groups.

#### Proactive

Proactive tasks, compared to steady-state walking, significantly modulate EMG-EMG coherence in the Beta and Gamma bands between synergistic muscles. Visually-guided walking increased TA-TA coherence in Alpha (46, 48) and Beta-Gamma bands for young and old adults (27, 28, 46, 48). Spedden et al. (2019) reported small effect size of task in late swing for Beta (ES = 0.05, η²p) and Gamma bands (ES = 0.04, η²p) (28). Obstacle avoidance task increased intra-muscular TA-TA coherence (ES = 1.06, large effect) and inter-muscular BF-ST coherence (ES = 0.7, Wilcoxon’s rB, large effect) in Beta band during swing (Table 3). During an auditory-guided task (*i.e.,* Rhythmic Auditory Stimulation) with swing phase perturbation (*i.e.,* resistance at the leg’s distal end), RF-RF coherence in Beta band decreased compared to walking with resistance alone (Table 3). Then, inter-muscular GM-SOL coherence increased in Gamma band during the stance of long step task (ES = 0.56, η²p) (16, 38) (Table 3). In contrast, two studies reported no significant modulation in TA-TA and GL-SOL coherence in Alpha (27) and Beta bands (28) during swing for visually-guided walking task (Table 3). One study did no show a significant change in GM-GL coherence for obstacle avoidance task (52). And another study observed no significant change in ST-BF coherence in late swing to early stance for auditory-guided walking task (34) (Table 3).

#### Proprioceptive

Proprioceptive-driven tasks, compared to steady-state walking, generally increase EMG-EMG coherence across frequency bands (Alpha, Beta, Gamma) (Table 3). High-heeled shoes increased Beta band coherence between proximal lower limb muscles in young adults (ES = 1.14–1.64, large effect) (Table 3) (37). Asymmetric walking speeds increased TA-TA Alpha band coherence in both slow (ES = 0.21, η²p) and fast legs (ES = 0.17, η²p) during swing in both young adults and elderly (45). Beta-Gamma coherence increased during swing for both legs, with higher Gamma coherence in the slow leg (22, 23, 45). Alpha band oscillations increased in gastrocnemius (GM-GL; ES = 0.11, η²p) and plantar flexor muscles (GM-SOL) for slow (ES = 0.21, η²p) and fast legs (ES = 0.18, η²p), with a moderate age-related effect in plantarflexors (ES = 0.12, η²p) (45). Beta-Gamma coherence increased only in the slow leg for GM-SOL coherence (ES = 0.08, η²p) (45) (Table 3). Abrupt swing phase perturbations increased TA-TA coherence in late swing (53) (Table 3). Nevertheless, some studies reported an absence of significant effect on EMG-EMG coherence. One study observed no significant change in Alpha band for asymmetric walking speeds in young adults (23). Another study reported no significant change in GM-GL coherence during stance in an abrupt, and gradual, swing phase perturbation tasks for Beta and Gamma bands (53). TA-TA coherence was not significantly changed by the gradual swing phase perturbation task during swing phase for Beta and Gamma bands (53) (Table 3).

#### Cognitive load

Dual-task affected EMG-EMG coherence according to age. Adding a cognitive task (*e.g.,* digit 2-back, watching figures) to locomotion did not affected significantly Beta and Gamma band TA-TA and GM-GL coherence in young adults (p > 0.05, ES < 0.39, small effect) (26) (Table 3). In older adults, Beta band TA-TA coherence increased in late swing during high (digit 2-back) vs. low (watching figures) cognitive load (p = 0.043, ES = 0.46, small effect) (26), while Gamma band GM-SOL coherence decreased in late stance during high cognitive load vs. a regular walk (16) (Table 3). Moreover, a lower intra-muscular and inter-muscular coherence during a choice reaction task compared to a simple reaction task in Alpha (p = 0.004) and Beta (p = 0.008) bands (21) for older adults (Table 3).

## DISCUSSION

### Results summary

This review aimed to summarize age and task-dependent modulations in EMG-EMG coherence during gait. Three main findings are worth highlighting. <u>First</u>, EMG-EMG coherence changes with age during regular walking, increasing in the Gamma band during maturation and decreasing in both the Beta and Gamma bands as part of the aging process. <u>Second</u>, EMG-EMG coherence is task-dependent in both young and older adults, showing significant increase in Alpha, Beta and Gamma bands during proactive and proprioceptive locomotor tasks in comparison to regular walk. However, young adults exhibit greater modulation of EMG-EMG coherence in the Beta and Gamma bands compared to older adults. <u>Third</u>, distinct EMG-EMG coherence responses seem to have emerged between young and older adults during proprioceptive-driven tasks and during walking with an additional cognitive load.

### Interpretation of EMG-EMG coherence

According to this review, the Beta band is the most studied frequency band and its significance seems to be well-known, however, the heterogeneity of interpretations of EMG-EMG coherence in Delta, Alpha, and Gamma bands makes our findings unclear and difficult to interpret.

#### Delta

The only study that clearly identifies the Delta frequency band did not interpret it in relation to the results during walking (41). The other study that included the corresponding Delta frequencies interprets these “low frequencies” coherence as representations of the afferent pathway during gait (40). This lack of studies makes the EMG-EMG coherence in the Delta frequency band difficult to interpret, particularly because this frequency range is commonly associated with movement artifacts (54).

#### Alpha

A consensus tends to emerge about the interpretation of the Alpha frequency band in this review. Several studies associate the increase in Alpha band coherence and the changes in movement coordination to a greater contribution of subcortical circuits and sensory feedback during gait (20, 44, 45, 50). However, the exact mechanisms related to this frequency band still unclear (55). Charalambous et al. (2022) assumed that the Alpha band is a proxy for the corticoreticulospinal tract and relies on different mechanisms than the Beta band, but the lack of studies investigating the direct association between this descending pathway and Alpha EMG-EMG coherence during gait makes this interpretation questionable (55). Additionally, as the corticoreticulospinal tract is mainly responsible for axial and proximal muscles during gross movements (55), but five studies observed intra-muscular coherence modulation in a distal lower limb muscle (TA), which is mainly influenced by the corticospinal tract (21, 45, 46, 48, 56). Therefore, despite a consensus emerging on the interpretation of the Alpha band, caution should be taken regarding the exact mechanisms underlying Alpha band modulation in healthy participants during gait.

#### Beta

For EMG-EMG coherence, our findings reported that Beta band coherence was commonly associated with corticospinal common drive between two synergistic muscles pairs to execute voluntary movements during gait (27, 28, 35, 50, 52, 53) and motor performances (28, 36, 45, 53). Furthermore, two studies had indicated that coherence in this frequency band was also sensitive to the increased cognitive demands associated with locomotor tasks (21, 34), even if one did not (26). These observations were in line with previous research that indicated that intra- and inter-muscular coherence were mainly driven by the motor cortex (57). Moreover, it is important to note that Norton and Gorassini (2006) reported an influence of corticospinal tract integrity on inter-muscular Beta band-associated frequency coherence (58). Therefore, the literature seems to have established a consensus on the corticospinal origins of Beta band coherence during gait using EMG-EMG coherence.

#### Gamma

With the Beta band, the EMG-EMG Gamma band coherence was widely used as a marker of common inputs originating from the corticospinal tract (16, 23, 27, 28, 38, 44, 45). Despite some studies using EEG/MEG-EMG coherence methods suggested that Gamma band could be more involved than the Beta band in dynamic muscle contraction movements or tasks that require more attentional resources (*i.e.,* visual and somatosensory information) (59, 60), no clear consensus had been reached regarding the distinct roles of the Beta and Gamma bands during locomotion in EMG-EMG coherence analysis in this review. Only one study in this review suggested a specific role of Gamma band coherence involved to accurately control the ankle (24).

### Effect of age on EMG-EMG coherence during gait

#### Maturation

The development of gait in children is marked by an increase in Gamma band EMG-EMG intra-muscular coherence for the TA muscle (24). Increase in Gamma band coherence correlated with reduced gait variability (24) and supports gait adaptations during critical periods of neural plasticity, such as between the ages of 4–6 and 7–9 (24, 61). The positive correlation between age and peak Gamma band coherence suggests that Gamma band oscillation is sensitive to age during walking tasks (24, 61). Additionally, EMG-EMG coherence has already demonstrated a developmentally dependent modulation in other tasks, such as dorsiflexion steady isometric contraction (TA-TA) (24) and thumb abduction (abductor pollicis longus - abductor pollicis brevis) (62), with Beta band coherence increasing with age. Thus, EMG-EMG coherence appears to be a relevant measure for studying the maturation of neural strategies involved in the control of gait. However, current research is limited to steady-state walking and the Gamma band, emphasizing the need for further investigation of motor control development across various locomotor tasks and frequency bands, from the acquisition to the maturity of walking.

#### Aging

Regarding aging effects, the findings of this review showed that all studies that compared young and older adults reported a significant reduction in Beta and Gamma EMG-EMG coherence during walking. These observations were observed in the TA during the swing phase (28, 45, 49), as well as between synergistic ankle (*i.e.,* GL-GM, GL-SOL, GM-SOL, TA-PL) and knee (*i.e.,* RF-VL) muscle pairs (35, 36, 45) during the early stance phase. The decrease observed in Beta and Gamma bands coherence for older adults seems to indicate a reduced corticospinal common drive to muscles with similar functions (63), probably due to structural alteration in corticospinal pathways with aging (64–66). These reductions have been associated to impaired gait performance, including slower walking speeds, greater step-to-step variability, low performance in visually guided tasks (28, 36, 45), reduced ankle and knee stabilization during the loading response of gait (67, 68), and reduced toe clearance during the swing phase, all increasing the risk of tripping or falling (67). Despite the lower coherence values, it was shown that older adults exhibited increased Beta band coherence under fatigue, in contrast to young adults who show no significant changes (35), interpreted as a compensatory strategy to maintain neural control of ankle muscles and preserve motor output. To conclude, a consensus has been reached that Beta-Gamma EMG-EMG coherence in synergistic lower limb muscles decreases with aging during gait.

### Effect of task on EMG-EMG coherence during gait

#### Steady-state

EMG-EMG coherences in the upper and lower limbs during walking are modulated to meet phase-specific demands of the gait cycle during regular walking, and this in Delta, Alpha, Beta, and Gamma frequency bands (14, 33, 41, 47, 50). During the swing phase, TA-TA coherence prevents foot drop and stabilizes the ankle during heel strike (14, 41, 67). In the stance phase, coherence between synergistic plantarflexor muscle pairs supports stability and push-off for propulsion (14, 47). Coherence between upper and lower limbs during swing (Delt, TA, SOL, RF) and stance (Delt, TA, SOL, BF) phases suggests a shared neural drive coordinating all four limbs (50), highlighting that the movement of the arms is not merely a passive response but an active component of locomotion (69). These findings seemed to report a consensus about the role of phase-specific neural drive modulation in ensuring efficient and coordinated locomotion. It should be noted that the low Beta coherence between antagonist muscle pairs (*i.e.,* TA and GL) would reflects minimal common drive, consistent with the reciprocal activation of ankle muscles during gait (70). After an experimentally induced fatigue, Dos Santos et al. (2020) observed modulations in EMG-EMG coherence, with distinct age- and phase-related adaptations in Beta band inter-muscular coherence during gait (mentioned in the section “Effect of age on EMG-EMG coherence during gait”) (35). It is important to note that, to our knowledge, this was the only study that analyzed the effect of fatigue on EMG-EMG coherence in the context of locomotion. Then, most studies revealed an absence of Gamma band significant changes in fast walking (16, 42, 48), except for one that observed an increase in inter-muscular coherence in synergistic lower limb muscles in “high frequencies” (40). However, lower frequency bands (particularly Alpha and Beta bands) were widely observed to be enhanced at higher walking speeds in both upper (42) and lower limbs (16, 36, 39, 40, 42, 48), supported by an association with transitions in upper-lower limb coordination (42). These findings suggest that a consensus seems to have been established about the specific role of Alpha and Beta bands in walking speed and associated changes in coordination, in contrast to higher frequencies (*i.e.,* Gamma band). Lastly, the findings of this review showed that gait initiation tasks lead to an increase in EMG-EMG coherence in the Alpha, Beta, and Gamma bands before movement onset. Nonetheless, only two studies investigated this locomotor task (21, 44), which makes it challenging to establish a clear consensus.

#### Proactive

Proactive tasks included: obstacles, targets, long step, and auditory guided walking task. In this review, voluntary and anticipatory adaptations of gait patterns were reported to increase the coherence in Beta and Gamma bands. This increase was reported between pairs of synergistic muscles involved in propulsion (16, 27, 38), adjustments of appropriate lower limb trajectories during the swing phase (52) and within the TA that ensures precise foot placement (27, 28, 46, 48, 52). Therefore, these findings indicate a pronounced increase in Beta and Gamma bands coherence between synergistic lower limb muscle pairs during proactive tasks, suggesting an increased cortical involvement during proactive tasks. However, one study regarding auditory-guided walking during a perturbation task observed a decreased Beta band coherence (34), indicating that the guidance modality, *i.e.,* rhythmic auditory *vs.* visual, may affect the cortical involvement to the locomotor task. Concerning Alpha band coherence, the literature is unclear for this category of locomotor task, with contrasting results (27, 46, 48) so that no consensus can be assumed in this frequency band regarding proactive tasks.

#### Proprioceptive

Proprioceptive tasks included: split belt, unilateral resistance, and high heeled shoes walking tasks. EMG-EMG coherence showed an increase in the Alpha, Beta, and Gamma bands, with more evidences in the Beta and Gamma bands (22, 23, 45, 53) than in the Alpha band (23, 45). These increased common inputs in the Beta and Gamma bands between synergistic and antagonistic lower limb muscle pairs may be required to maintain temporal gait symmetry (*i.e.,* double support time, swing phase duration) (23, 34, 45), prevent imbalance around lower limbs (37), and, therefore, improve gait stability during gait perturbations. Moreover, larger gait perturbation increased EMG-EMG coherence modulation (53, 71, 72). These findings commonly highlighted the role of Beta and Gamma bands coherence, suggesting an increased cortical involvement during proprioceptive tasks. Interestingly, one study highlighted that although older adults showed less increase in the Beta-Gamma band during proprioceptive tasks than young adults. Alternatively, the increase of coherence was higher in older adult than in young adult in the Alpha band, accompanied by slower gait adaptations (45). In addition, the only two studies that reported Alpha band coherence had contrasting results. One study showed no modulation in intra-muscular coherence (23), while the other study showed an increase between plantarflexor muscle pairs (45). However, EMG-EMG coherence responses between ages and the involvement of Alpha band coherence in this type of locomotor task remain unclear.

#### Cognitive load

Cognitive load tasks included: auditory 2-back task, watching figures, digit 2-back task during a walking task, and a choice reaction task to initiate gait. The effect of dual-task on EMG-EMG coherence revealed complex interactions between cognitive load and motor control strategies. In the present review, three studies were included that investigated the effects of cognitive demands during locomotor tasks on EMG-EMG coherence (16, 21, 26). Two studies reported a decrease in intra- and inter-muscular coherence, one in the Alpha and Beta frequency bands (21) and the other in Gamma frequency band (16). These findings highlighted a tendency toward EMG-EMG coherence decrease during cognitive-motor task in older adults. In contrast, Kitatani et al. (2023) observed no significant change in intra-muscular (TA-TA) coherence in the Beta band during a high-cognitive-load dual-task in young adults (26). Thus, the age-related effect of cognitive load on EMG-EMG coherence is still unclear.

### Methodological considerations

Since surface EMG is influenced by both physiological and non-physiological sources of variability (73, 74), EMG-EMG coherence findings should be interpreted with caution. Regarding EMG signal processing, heterogeneity was observed across the included studies. First, most studies rectified EMG signals while its relevance is debated because it is considered less effective than raw EMG (referred to as “interference EMG”) in preserving frequency content (75, 76). Second, most studies used Fourier transforms to calculate coherence, however, wavelet-based coherence is particularly suited for non-stationary signals, making it particularly relevant for EMG-EMG coherence measurement during gait (77). Third, limitations are evident in frequency band analysis (55). Several studies computed coherence within frequency bands that were initially filtered during the preprocessing steps (23, 40–42, 46, 48, 49, 78). Future studies should carefully preserve the frequency content subsequently analyzed. Also, the frequency range of Alpha, Beta, and Gamma bands varied between studies and showed inconsistencies. For instance, two studies defined Alpha band as the 5-15 Hz frequency band, although 5-8 Hz is theoretically considered the Theta band (27, 47). Overlaps were reported between the Alpha and Beta bands whereas these frequency bands represent distinct neural processes. In some cases, frequency bands were very narrow to provide meaningful interpretations (*e.g.,* 36-40 Hz for the Gamma band in Gennaro & de Bruin 2020 (49)). As the literature assumes that EMG-EMG common drive is results from cortical and subcortical sources influences (79), it may be relevant to use the same frequency bands as described in magneto/electroencephalography analysis, namely, 8-13 Hz, 13-31 Hz and >31 Hz for Alpha, Beta, and Gamma bands, respectively. Only two studies analyzed the Delta band or associated frequencies (40, 41), and one study investigated the Theta band (42). On the one hand, this review highlights the lack of knowledge regarding these low-frequency bands EMG-EMG coherence during walking tasks and emphasizes the need to explore these bands under dynamic conditions. On the other hand, the interpretation of these frequency bands should be taken with caution due to movement artifacts that may occur within these frequency bands during gait as discussed in the section “EMG-EMG coherence interpretation”. Lastly, most studies reported only one frequency band, which could imply a selection bias. To ensure a better understanding of EMG-EMG coherence, all frequency bands should be investigated and reported in future investigations. Fourth, two studies in this review discussed the reliability of EMG-EMG measurements during walking (48, 49). Despite both studies analyzing the test-retest reliability of TA-TA coherence during the swing phase, their findings were contradictory. One study reported good to very good reliability (48), while the other indicated low reliability (49). To our knowledge, another study that was not included in this review assessed the reliability of EMG-EMG measurements during walking (TA-TA) and highlighted the dependence on specific conditions (*e.g.,* slow walking speed, less task complexity) and EMG settings (*e.g.,* rectified signal and high-pass with 10 Hz) for qualifying EMG-EMG coherence as a reliable method (80). The significant influence of experimental conditions limits the practical use of this method, and EMG settings should be carefully considered when assessing significant modulation in coherence. Therefore, no consensus emerged from the current literature regarding EMG signal processing. Future investigations should aim to establish clear guidelines for EMG-EMG coherence analysis during walking to enhance the relevance, interpretation, and comparability of findings across studies.

Another important aspect to consider is that treadmill walking accounted for 74% of the walking conditions among the included studies. Although it allows the study of gait in a controlled environment and ensuring a consistent number of gait cycles, treadmill walking involves different corticospinal motor control mechanisms compared to overground walking, which better reflects ecological condition (68). Indeed, treadmill walking reduces kinematic variability and decreases the need for active regulation of spatiotemporal gait parameters (81, 82). In contrast, overground walking requires dynamic and autonomous adjustments of speed and stride, which implies greater demands on cortical control and corticospinal interactions (68). To our knowledge, no studies have mentioned a direct comparison between overground and treadmill walking, but these differences may affect EMG-EMG coherence by reducing coherence in higher frequency bands (*i.e.,* Beta and Gamma). Therefore, findings from treadmill conditions should be interpreted with consideration of these reduced locomotor demands, and further studies should explore EMG-EMG coherence in more ecological conditions.

### Limitations

The limitations of this scoping review must be acknowledged. While analyzing kinematics in relation to changes in EMG-EMG coherence would have been interesting, direct associations were not included in this review. Although nearly all the studies analyzed kinematic parameters, such as gait variability or spatiotemporal parameters of walking (with only four exceptions (14, 21, 49, 51)), there was a risk that the primary variable of interest (EMG-EMG coherence) might have been obscured by the extensive array of results. The same reasoning applies to associations with directionality (44) and men/women distinction.

## CONCLUSION

This scoping review highlighted the age- and task-modulation of EMG-EMG coherence during gait. Findings reported a consensus on the decrease in EMG-EMG coherence during walking with aging, particularly in the Beta and Gamma bands, which could be attributed to alterations in the corticospinal tract with aging. Moreover, Beta and Gamma EMG-EMG coherence tended to increase during challenging proprioceptive and proactive locomotor tasks, which is interpreted as an increase in cortical involvement to control the gait pattern. The results of this review also emphasize the need for future research to investigate EMG-EMG coherence in other frequency bands, such as the Alpha band, using standardized signal processing methods and frequency classifications to ensure a better understanding of EMG-EMG coherence in locomotor tasks, and to explore coherence in children across various locomotor tasks to better understand its association with maturation.

## APPENDIX

### Appendix 1

Database(s): **Ovid MEDLINE(R) ALL** 1946 to March 28, 2024

Search Strategy:

| # | Searches |
| --- | --- |
| 1 | (coherence? or synerg* or coordination or "activation pattern" or "activation patterns" or "nonnegative matrix factorization" or "nonnegative matrix factorisation" or "common drive" or "neuromuscular control" or "motor control" or "principal component analysis").ab,kf,ti. |
| 2 | (muscle? or muscular or intermuscular or corticomuscular or electromyograph* or electromyogram*).ab,kf,ti. |
| 3 | Electromyography/ or Muscle, Skeletal/ |
| 4 | 2 or 3 |
| 5 | ("lower limb" or "lower limbs" or "lower extremity" or "lower extremities" or leg or legs).ab,kf,ti. |
| 6 | exp Lower Extremity/ |
| 7 | 5 or 6 |
| 8 | (gait or walk* or locomotion).ab,kf,ti. |
| 9 | locomotion/ or walking/ or gait/ |
| 10 | 8 or 9 |
| 11 | 1 and 4 and 7 and 10 |
| 12 | exp Animals/ not (exp Animals/ and Humans/) |
| 13 | 11 not 12 |

## DATA AVAILABILITY

Data are available upon request to the corresponding author.

## ACKNOWLEDGEMENTS

The authors would like to thank Sina Tabeiy and Sina Esmaeili for their contribution to the screening of the articles.

## AUTHORS CONTRIBUTIONS

**S.D.** analyzed and interpreted results of studies, prepared figures/tables, and drafted manuscript; **Y.C.** contributed to study conceptualization, research strategy development, the screening process, result interpretations, supervision, and major revision of the manuscript. **D.A.** co-conceived and designed the research strategies. **F.D.M.** was involved in result interpretations, supervision and major revisions of the manuscript. **L.F.** was also involved in result interpretations and the revision of the final version of the manuscript. All authors contributed to the writing, review, and editing of the manuscript.

